# Deciphering autoantibody landscape of systemic sclerosis through systems-based approach: insights from a B-cell depletion clinical trial

**DOI:** 10.1101/2024.07.30.24311212

**Authors:** Kazuki M Matsuda, Satoshi Ebata, Kazuhiro Iwadoh, Hirohito Kotani, Teruyoshi Hisamoto, Ai Kuzumi, Takemichi Fukasawa, Asako Yoshizaki-Ogawa, Shinichi Sato, Ayumi Yoshizaki

## Abstract

Systemic sclerosis (SSc) is a progressive fibrotic disorder with a high mortality rate, characterized by extensive autoantibody production. Despite recent advancements, effective treatments remain limited. Rituximab (RTX), a B-cell depleting agent, has shown promise in clinical trials. The DESIRES trial highlighted the reduction in modified Rodnan Skin Score (mRSS) and the association between serum immunoglobulin levels and RTX responsiveness. We employed proteome-wide autoantibody screening (PWAS) using wet protein arrays (WPAs) that display 13,455 human autoantigens to analyze serum samples from SSc patients in the DESIRES trial and age- and sex-matched healthy controls (HCs). As a result, the sum of autoantibody levels (SAL) was significantly higher in SSc patients compared to HCs. High responders (HRs) to RTX showed a greater initial SAL and significant reductions post-treatment, unlike low responders (LRs). Machine learning identified specific autoantibodies linked to disease status, and 58 autoantibodies were identified as clinically relevant. Some of those autoantibodies targeted membrane proteins including G protein-coupled receptors, associated with better differentiation between HRs and LRs. Our findings underscore the significance of autoantibodies in SSc pathogenesis and their potential role in predicting RTX responsiveness. This comprehensive autoantibody profiling could enhance diagnostic and therapeutic strategies, and moreover, better understanding of the pathophysiology of SSc.

## Introduction

Systemic sclerosis (SSc) is a progressive fibrotic disorder that affects both the skin and internal organs.^1^ Among connective tissue diseases, it has the poorest prognosis, with an approximate 30% mortality rate within ten years.^2^ Historically, treatment options for SSc have been limited. While the exact cause of SSc remains unclear, increasing evidence indicates that B cells play a significant role in the pathogenesis.^3–5^ They contribute by producing autoantibodies, secreting unique cytokines, and activating other immune cells. Reflecting this understanding, various B-cell targeting therapeutic modalities, monoclonal antibodies,^6–8^ Bruton’s tyrosine kinase inhibitors,^9^ and chimeric antigen receptor (CAR) T-cell therapies,^10–12^ are emerging as promising treatments for SSc.

Rituximab (RTX) is a chimeric monoclonal antibody that depletes circulating B cells by targeting the B-cell specific antigen CD20. Previously, we conducted a double-blind, investigator-initiated, randomized, placebo-controlled trial, or the DESIRES trial,^13^ which demonstrated significant superiority of RTX over placebo by the absolute reduction of modified Rodnan Skin Score (mRSS) 24 weeks after initiation of the study period. The open-label extension of this trial also revealed the long-term efficacy and safety of RTX on SSc,^14^ as well as the association between decrease in serum immunoglobulins and greater clinical response to RTX.^15^

The emergence of autoantibodies is a prominent feature of SSc, highlighting its nature as an autoimmune disorder. Anti-nuclear antibodies (ANAs), detected through indirect immunofluorescence on HEp-2 cells, are present in more than 90% of patients with SSc.^16^ Several ANAs are specific to SSc or are closely associated with distinct clinical subsets. For instance, anti-topoisomerase I antibodies (ATA) are strongly linked to diffuse skin sclerosis and severe interstitial lung disease (ILD), while anti-centromere antibodies (ACA) are primarily associated with limited skin and lung involvement.^17^ However, the pathogenicity of these ANAs is debated, as they cannot reach their targets across the plasma and nuclear membranes *in vivo*. Additionally, the DESIRES trials revealed that serum ATA levels did not decrease,^13,14^ indicating that the relationship between serum immunoglobulin levels and responsiveness to RTX cannot be explained by ATA levels.

Herein, we employed our original technique for proteome-wide autoantibody screening (PWAS) using wet protein arrays (WPAs) that cover approximately 90% of the human transcriptome.^18,19^ This technique has previously been used to develop multiplex measurements for disease-related autoantibodies,^20,21^ identify clinically relevant novel autoantibodies,^22,23^ and investigate inter- and intra-molecular epitope spreading during the disease course. We applied PWAS to serum samples from SSc patients who participated in the DESIRES trial, as well as sex- and age-matched healthy controls (HCs). Our aim was to elucidate the autoantibody landscape in SSc and investigate clusters of autoantibodies that contribute to the reduction of serum immunoglobulin levels associated with a good response to RTX in SSc patients. This research seeks to identify novel biomarkers and gain a better understanding of the pathogenesis of SSc.

## Results

### Subjects

In the DESIRES trial (NCT04274257), a total of 56 individuals diagnosed with SSc were evenly randomized into two groups: 28 received RTX and 28 received a placebo.^13^ Serum samples were collected at the start and after 24 weeks of treatment from participants who completed the term. These samples were reacted with WPAs for PWAS (**Figure 1A**). After excluding samples that were not suitable for PWAS, such as those containing anti-GST-tag antibodies, the study proceeded with 24 patients in the RTX group and 21 patients in the placebo group. The baseline characteristics of the two groups were comparable (**Table 1**). An equal number of age- and sex-matched healthy controls (HCs) were also included in the study (**Figure 1B**).

**Figure 1.**
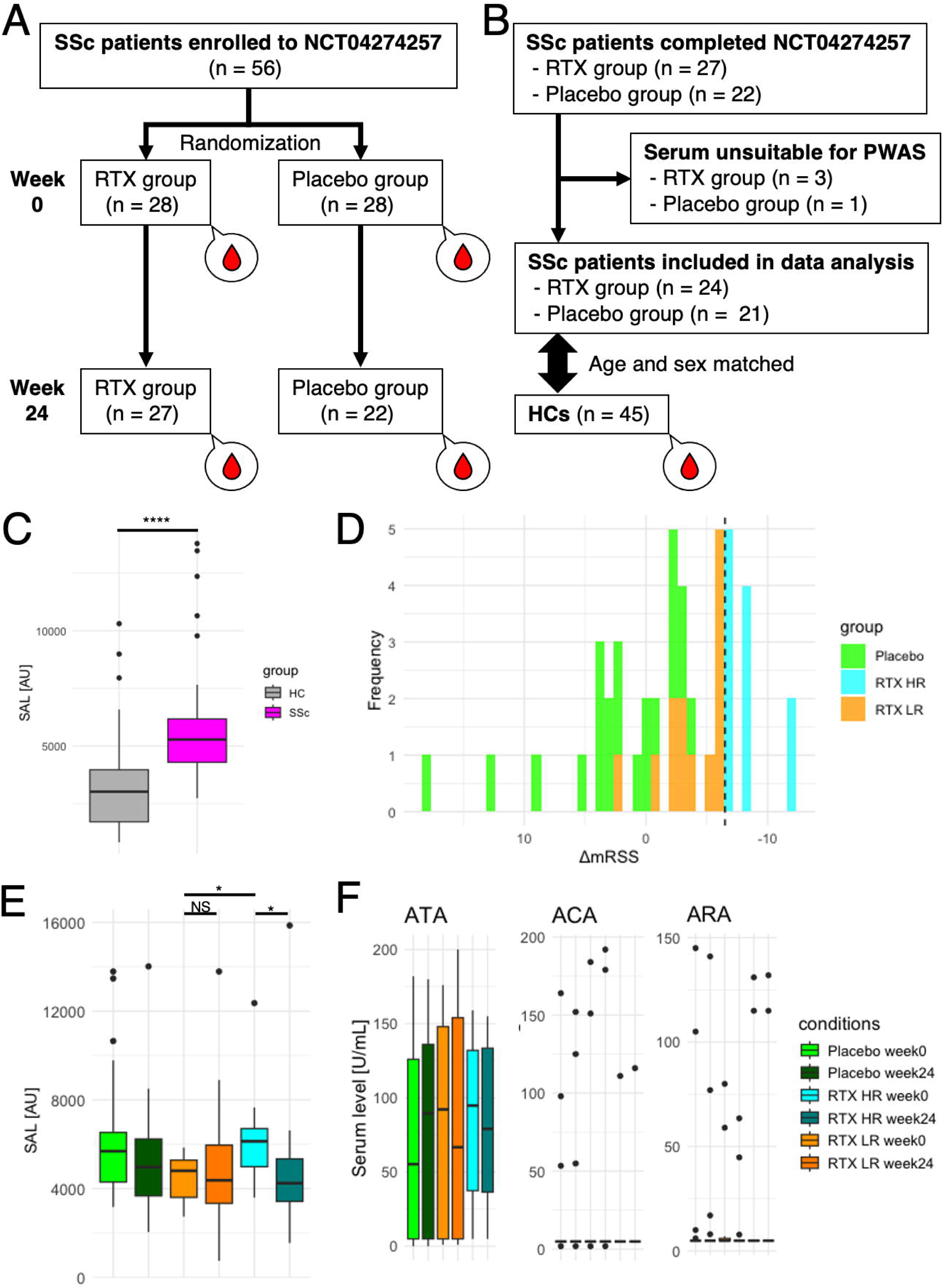
Overview of the study. **(A)** The study design of the DESIRES trial. **(B)** The flow chart of sample acquisition in the present study. (C) The sum of autoantibody levels (SAL) in SSc at the baseline and HCs. (D) Absolute change of mRSS in SSc patients during the DESIREs trial. HR: high responder, LR: low responder. (E) SAL before and after the study period of the DESIRES trial by the treatment arm. (F) Serum levels of SSc-related autoantibodies before and after the study period of the DESIRES trial by the treatment arm. ATA: anti-topoisomerase I antibody, ACA: anti-centromere antibody, ARA: anti-RNA polymerase III antibody. *: P < 0.05, ****: P < 0.0001, NS: P > 0.05. P values were calculated by Mann-Whitney’s U test.

### The sum of autoantibody levels

We defined the sum of autoantibody levels (SAL) as the total serum concentration of all autoantibodies assessed in our PWAS. SAL was significantly higher in SSc patients compared to HCs at the baseline (P < 0.0001; **Figure 1C**). Subsequently, we categorized the RTX treatment group into two subgroups based on their response to treatment (**Figure 1D**): high responders (HRs; n = 11) and low responders (LRs; n = 13). Initially, SAL was significantly greater in the HR group compared to the LR group (P < 0.05; **Figure 1E**). Additionally, SAL levels decreased significantly from week 0 to week 24 in the HR group (P < 0.05), whereas changes in the LR group were not statistically significant. This pattern persisted across all age groups (**Extended Figure 1A**). In contrast, serum levels of well-known SSc-related autoantibodies, including anti-topoisomerase I antibodies (ATA), anti-centromere antibodies (ACA), and anti-RNA polymerase III antibodies (ARA), did not follow this trend, as observed both in clinical-standard ELISA tests (**Figure 1F**) and in our PWAS (**Extended Figure 1B**).

### Machine learning

To determine which autoantibodies were driving the association between the SAL and disease status, we firstly utilized nine machine learning frameworks. Notably, Lasso regression, Ridge regression, SVM with normalization, XGBoost, and LightGBM achieved an area under the receiver-operator characteristics curve (ROC-AUC) exceeding 0.96, demonstrating an almost perfect ability to distinguish between SSc patients and HCs (**Table 2**). We identified the top 10 features in these six models (**Extended Figure 2A**), examined their inclusion relationships (**Extended Figure 2B**), and explored the prevalence of highlighted autoantibodies across a broader range of human disorders using the aUToAntiBody Comprehensive Database (UT-ABCD).^24^ Most of these autoantibodies were found to be non-specifically elevated in various pathological conditions, except for well-known SSc-specific autoantibodies such as ATA and ARA (**Extended Figure 2C**). Attempts to distinguish between HRs and LRs based on pre-treatment autoantibody profiles did not yield satisfactory precision (**Extended Table 1**).

**Figure 2.**
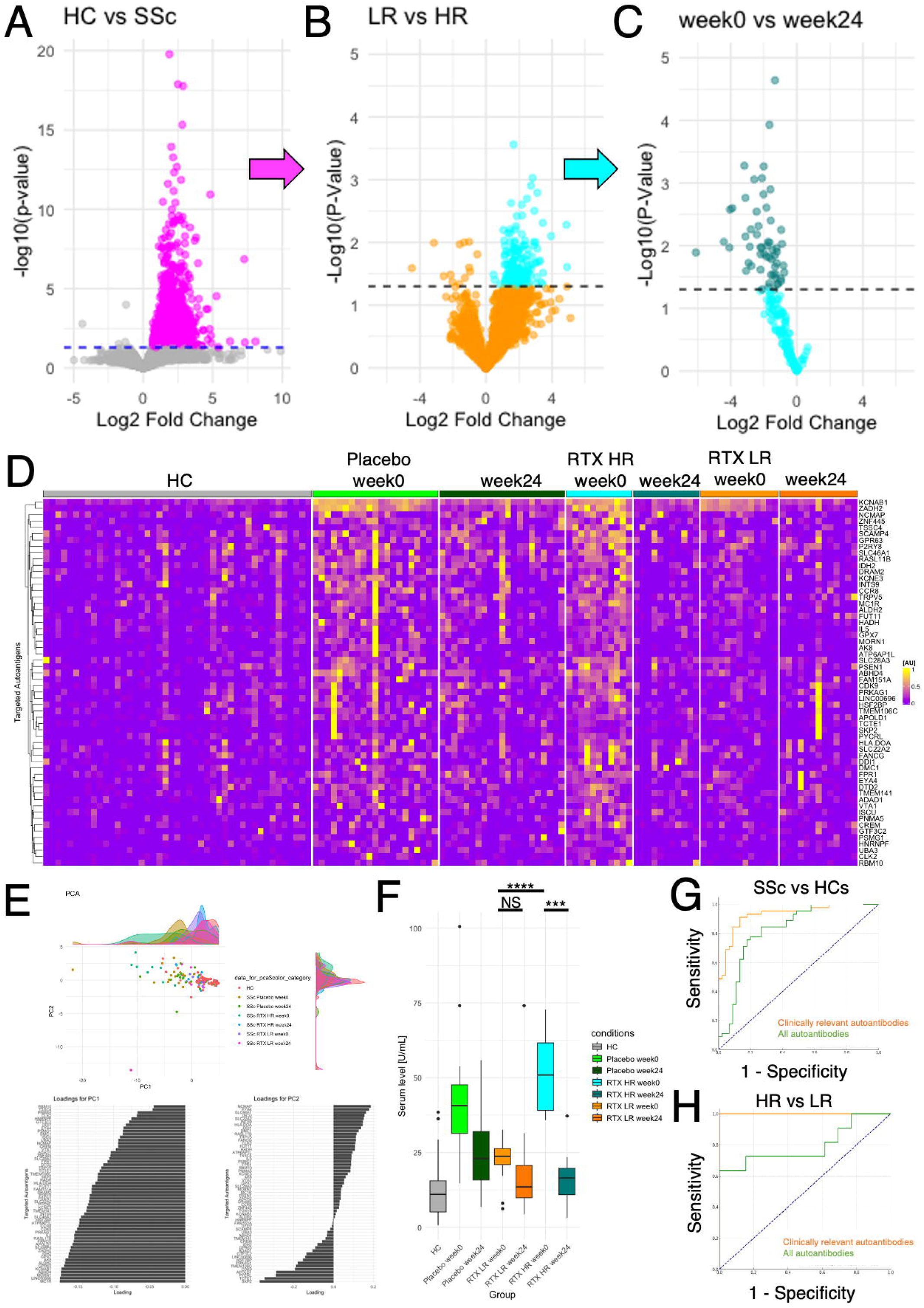
Selection of autoantibodies of clinical relevance. **(A)** Volcano plot that shows autoantibodies differentially elevated in SSc before treatment compared to HCs. The vertical dash line indicates P = 0.05. **(B)** Volcano plot that shows autoantibodies differentially increased in HRs than in LRs. **(C)** Volcano plot that shows autoantibodies significantly reduced in HRs during RTX therapy. **(D)** Heat map that shows the serum levels of 58 candidate autoantibodies of clinical relevance. **(E)** Principal component analysis of 58 candidate autoantibodies of clinical relevance. In the scatter plot, individual subjects as points. The loading diagram illustrates the contributions to PC1 and PC2. **(F)** SAL focused on 58 candidate autoantibodies of clinical relevance. ***: P < 0.001, ****: P < 0.0001, NS: P > 0.05. P values were calculated by Mann-Whitney’s U test. **(G)** The receiver-operator characteristics (ROC) curve demonstrating the discrimination between SSc and HCs by all the measured autoantibodies (green) and by 58 candidate autoantibodies of clinical relevance (yellow). **(H)** The ROC curve demonstrating the discrimination between HRs and LRs by all the measured autoantibodies (green) and by 58 candidate autoantibodies of clinical relevance (yellow).

### Investigating clinically relevant autoantibodies

We next sought to identify autoantibodies significantly associated with the clinical features of SSc using the trial data that included longitudinal changes. Initially, we selected autoantibodies that were significantly elevated in SSc patients compared to HCs at baseline (**Figure 2A**). Next, we chose autoantibodies that were significantly higher in HRs compared to LRs (**Figure 2B**). Lastly, we focused on autoantibodies whose serum concentrations significantly decreased from week 0 to week 24 in HRs (**Figure 2C**). As a result, 58 autoantibodies were identified as candidates for clinically relevant autoantibodies in SSc (**Figure 2D**). Principal component analysis (PCA) indicated that these 58 autoantibodies appear to distinguish HRs at week 0 within the dataset (**Figure 2E**). The sum of the serum levels of these autoantibodies displayed a pattern like SAL, but in a more pronounced manner (**Figure 2F**). Consistently, ROC-AUC analysis showed that these 58 autoantibodies provide better differentiation between SSc patients and HCs compared to the complete autoantibody profile (**Figure 2G**), and moreover, distinguish between HRs and LRs with nearly perfect precision (**Figure 2H**).

### Profiling candidate autoantibodies

To explore the molecular features of the human proteins targeted by the identified candidate autoantibodies, we conducted gene ontology enrichment analyses utilizing Metascape.^25^ Notably, whereas established SSc-related autoantibodies primarily target intracellular antigens, our findings emphasized membrane proteins associated with processes such as “import across plasma membrane” and “peptide G protein-coupled receptors (GPCRs)” (**Figure 3A**). There were four autoantibodies linked to each of the processes “import across plasma membrane” and “peptide GPCRs” (**Figure 3B**). Autoantibodies associated with plasma membrane import included those targeting presenilin 1 (PSEN1), solute carrier family 22 member 2 (SLC22A2), transient receptor potential cation channel subfamily V member 5 (TRPV5), and solute carrier family 46 member 1 (SLC46A1; **Figure 3C**). The group related to peptide GPCRs comprised autoantibodies against chemokine receptor 8 (CCR8), formyl peptide receptor 1 (FPR1), melanocortin 1 receptor (MC1R), and G protein-coupled receptor 63 (GPR63; **Figure 3E**). The associations between each autoantibody and the clinical characteristics of SSc were detailed in **Figures 3D and 3F**, which highlighted positive correlation between anti-TRPV5 and anti-CCR8 antibody levels and mRSS, as well as negative correlation between anti-FPR1 antibody levels and forced lung capacity. We also investigated the distribution of these highlighted autoantibodies in a broader population, utilizing UT-ABCD. As a result, all the items were not specific to SSc (**Extended Figure 3**).

**Figure 3.**
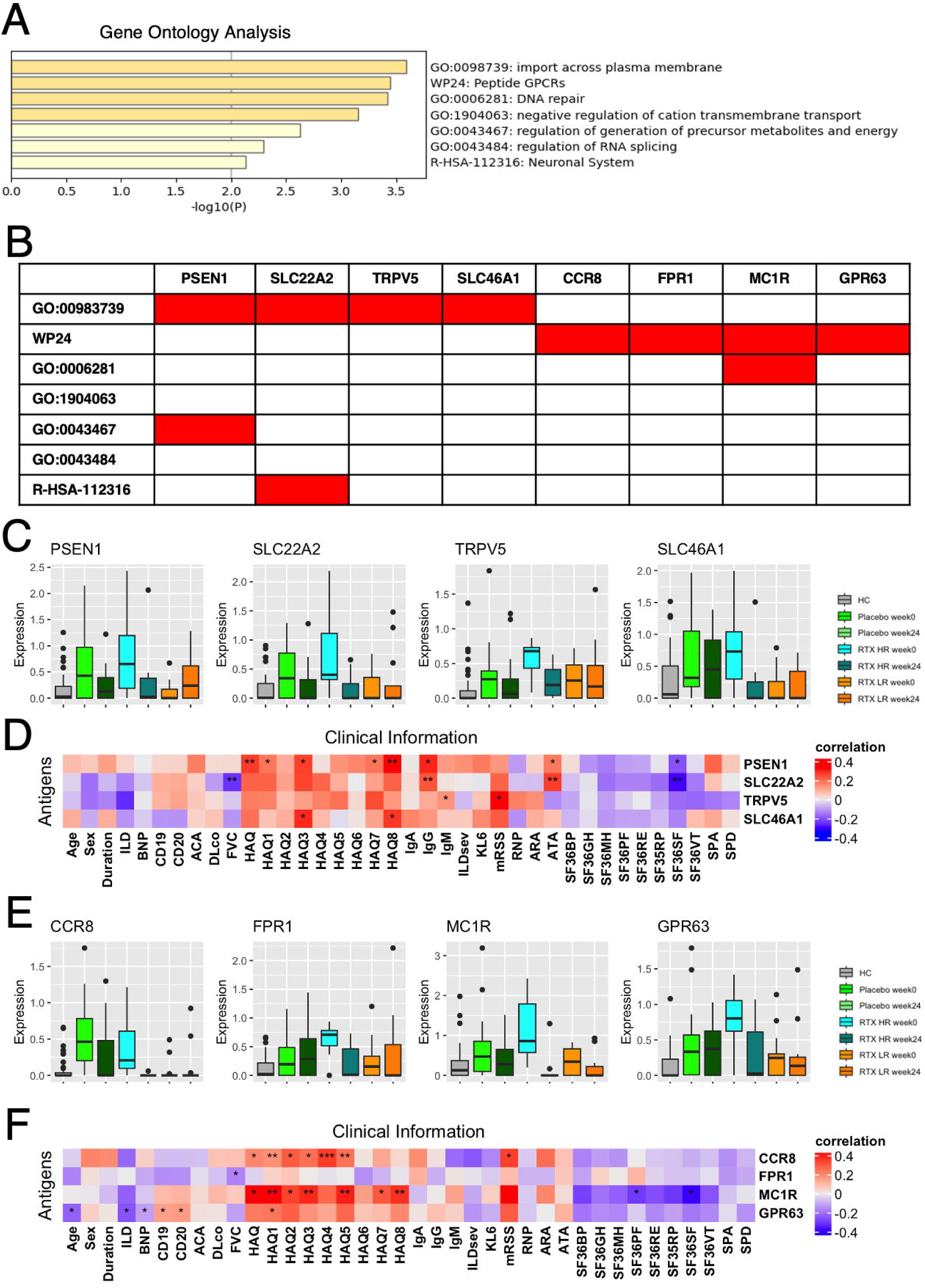
Proteins targeted by the candidate autoantibodies of clinical relevance. **(A)** Gene ontology analysis encompassing the genes coding proteins targeted the candidate autoantibodies. **(B)** The table represents the inclusion relationship between each gene ontology and the candidate autoantibodies. **(C)** Serum levels of candidate autoantibodies associated with “import across plasma membrane.” **(D)** The heatmap illustrates correlation between the candidate autoantibodies associated with “import across plasma membrane” and clinical traits of SSc. **(E)** Serum levels of autoantibodies associated with “peptide GPCRs.” **(F)** The heatmap illustrates correlation between the candidate autoantibodies associated with “peptide GPCRs” and clinical traits of SSc. *: P < 0.05, **: P < 0.01, ***: P < 0.001. P values were calculated by Spearman’s correlation test.

The tissue specificity of the highlighted autoantigens was investigated using publicly available databases. Analysis of multiple human tissues through bulk RNA sequencing data from the Human Protein Atlas^26^ revealed that *CCR8* and *FPR1* are enriched in bone marrow and lymphoid tissues (**Extended Figure 4, A and B**). Single-cell RNA sequencing data from the Tabula Sapiens project^27^ showed that *CCR8* expression is enhanced in regulatory T cells (Tregs; **Extended Figure 4C**), while *FPR1* expression is elevated in neutrophils (**Extended Figure 4D**). Meanwhile, *TRPV5* expression was localized in the kidney, and *MC1R* expression appeared to be relatively ubiquitous across different tissues.

**Figure 4.**
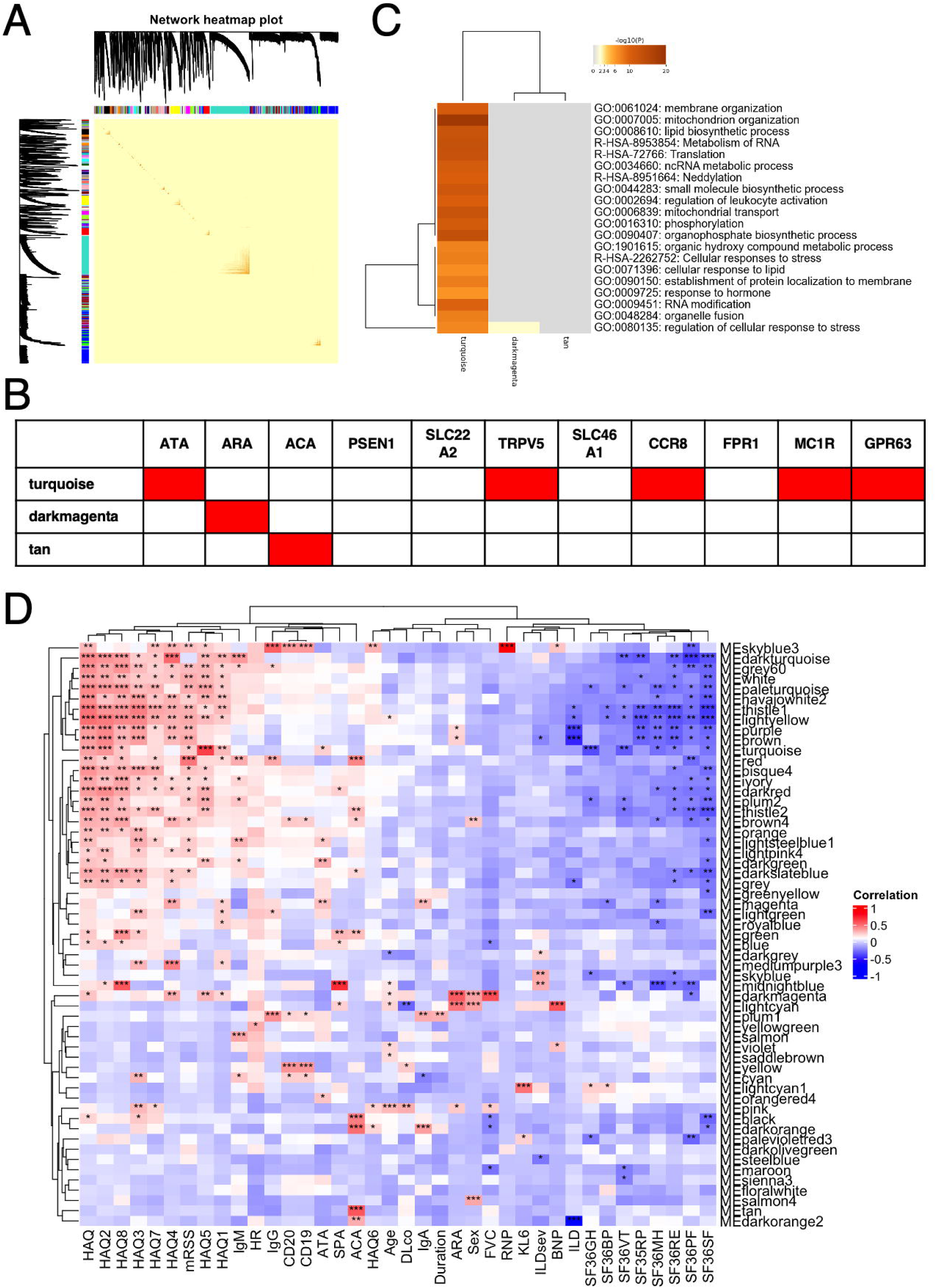
WGCNA analysis. **(A)** Network heatmap plot. Branches in the hierarchical clustering dendrograms correspond to modules. Color-coded module membership is displayed in the color bars below and to the right of the dendrograms. In the heatmap, high co-expression interconnectedness is indicated by progressively more saturated yellow and red colors. **(B)** The table represents the inclusion relationship between autoantibodies and modules identified by WGCNA analysis. **(C)** Gene ontology analysis encompassing the genes coding proteins targeted by autoantibodies included in the “turquoise,” “darkmagenta,” and “tan” modules. **(D)** The heatmap shows the correlation between each module and clinical trait. *: P < 0.05, **: P < 0.01, ***: P < 0.001. P values were calculated by Spearman’s correlation test.

### Weighted correlation network analysis

We utilized weighted correlated network analysis (WGCNA)^28^ to delve deeper into the correlations between autoantibodies in SSc. Our study included 135 specimens from SSc patients at weeks 0 and 24, as well as HCs. We constructed a correlation network for all autoantibodies evaluated in our PWAS and identified 57 distinct modules (**Figure 4A**). The “turquoise” module, which contained ATA, also included half of the candidate autoantibodies: anti-TRPV5, CCR8, MC1R, and GPR63 antibodies (**Figure 4B**). The “darkmagenta” module included ARA. The “tan” module included ACA. The other candidate autoantibodies did not appear in either the “darkmagenta” or “tan” modules. We also conducted gene ontology analyses on the gene lists encoding proteins targeted by autoantibodies within these three modules (**Extended Table 2**). The Enrichment score was the highest for “mitochondrion organization” in the “turquoise” module (**Figure 4C)**. The enrichment scores for “regulation of cellular response to stress” were highest in the “turquoise” module, moderate in the “darkmagenta” module, and lowest in the “tan” module. This pattern aligns with the clinical severity associated with ATA, ARA, and ACA.

We also reviewed the associations between each module and clinical traits (**Figure 4D)**. The “turquoise” module was positively linked to ATA, a higher modified Rodnan Skin Score (mRSS) – the primary endpoint of the DESIRES trial – as well as to lower patient-reported quality of life, evidenced by higher Health Assessment Questionnaire (HAQ) scores and lower 36-Item Short Form Health Survey (SF-36) scores. The “darkmagenta” module was associated with ARA positivity and higher forced vital capacity of the lungs, indicating a lower degree of ILD. The “tan” module was correlated with the ACA profile but did not show any significant associations with other clinical traits

## Discussion

In this study, we applied our original PWAS technique subjecting serum samples from SSc patients who participated in the DESIRES trial, along with sex and age-matched HCs. Our findings revealed a significant elevation in the overall amount of autoantibodies in SSc patients compared to HCs (**Figure 1**). We identified 58 autoantibodies as clinically relevant candidates in SSc, which showed a strong response to B-cell depletion therapy (**Figure 2**). Gene ontology analysis highlighted autoantibodies targeting membranous proteins, including transmembrane transporters and GPCRs (**Figure 3**). Most of them were clustered in the same module identified by WGCNA, which was significantly associated with ATA positivity, higher mRSS, and lower patient-reported quality of life **(Figure 4**). We demonstrated the nearly perfect distinction between SSc and HC samples achieved through machine learning (**Table 2**), and SSc patients who show good response to RTX treatment engaging the clinically relevant autoantibodies highlighted in our analyses (**Figure 2H**). These results underscore the effectiveness of our integrating systems-based comprehensive autoantibody screening and bioinformatic analyses, which could be called as “autoantigenomics.“^29^

Our machine learning analysis confirmed the prominence of well-known SSc-specific ANAs such as ATA and ARA (**Extended Figure 2**). However, these autoantibodies did not exhibit longitudinal changes during the clinical trial nor differences between HRs and LRs to RTX treatment (**Figure 1F and Extended Figure 1B**), consistent with our previous findings.^13,14^ In contrast, we observed that changes in the overall levels of autoantibodies were significantly correlated with RTX efficacy (**Figure 1, C and E**). This discrepancy led us to investigate autoantibodies that exhibit significant associations with the time course and drug response to RTX therapy. While predictive factors for RTX responsiveness have been limited to clinical indicators such as CD19-positive cell counts, mRSS, and serum levels of SP-D,^30^ the clinically relevant autoantibodies we identified were able to discriminate between HRs and LRs with high accuracy (**Figure 2H**). Interestingly, some of these autoantibodies targeted membranous antigens (**Figure 3**), suggesting that they are not only useful as biomarkers but also could play a direct role in the pathogenesis of SSc.

Accumulating evidence has unveiled the relevance of autoantibodies targeting GPCRs in immune-related human disorders, including SSc.^31,32^ Notably, previous studies have indicated the relationship between GPCRs targeted by the candidate autoantibodies and the pathogenesis of SSc. CCR8 is primarily expressed on Tregs (**Extended Figure 4C**), which suppress autoimmune responses.^33^ CC Chemokine 1 (CCL1) acts as a chemoattractant, recruiting Tregs to peripheral tissues by binding to CCR8.^34^ While direct evidence linking the CCR8-CCL1 axis to SSc pathogenesis is lacking, blocking CCR8 could exacerbate autoimmune responses in SSc by inhibiting Treg homing to target tissues. FPR1, which is abundantly expressed on neutrophils (**Extended Figure 4D**), induces inflammation when bound by its cognate ligands.^35^ One such endogenous agonist is N-formyl-methionine (fMet), a mitochondrial protein-derived molecule that serves as a damage-associated molecular pattern.^36^ Lurley R et al. have shown in SSc, elevated serum fMet levels and fMet-induced activation of neutrophils through FPR1-dependent mechanisms.^37^ Consistent with these findings, our enrichment analysis highlighted mitochondrion-associated antigens targeted by autoantibodies in SSc (**Figure 4C**), suggesting mitochondrial destruction, which has also been reported in other autoimmune conditions such as systemic lupus erythematosus and rheumatoid arthritis. MC1R is expressed on various cell types (**Extended Figure 4B**), including melanocytes, inflammatory cells, endothelial cells,^38^ and skin fibroblasts from SSc patients.^39^ Its endogenous ligand, α-melanocyte-stimulating hormone, induces melanin production in melanocytes and promotes anti-inflammatory responses, such as inhibition of nuclear factor-κB and suppression of pro-inflammatory cytokines.^40^ Kondo M et al. demonstrated that dersimelagon (MT-7117), an oral MC1R agonist, has favorable effects on inflammation, vascular dysfunction, and fibrosis, which are key pathologies in preclinical SSc models.^41^ This prompted a phase 2 clinical trial to evaluate the efficacy and tolerability of MT-7117 in patients with early, progressive diffuse cutaneous SSc (NCT04440592). Collectively, accumulating experimental and clinical evidence suggests that modulating these GPCRs, either through stimulation or blockade, could potentially alter SSc disease severity.

Meanwhile, it is important to note that the serum levels of the candidate autoantibodies we identified were relatively modest compared to conventional SSc-specific autoantibodies like ATA (**Extended Figure 1B**). Moreover, the presence of these candidate autoantibodies did not appear to be specific to SSc, as indicated by data from UT-ABCD (**Extended Figure 3A and 3B**). These suggest that the pathogenesis of SSc may not be attributed to a single unique autoantibody that shows strong cross-reactivity with conventional SSc-specific autoantibodies, but rather to a collaboration of multiple autoantibodies. This hypothesis aligns with several observations that suggest that SSc is a multifactorial disease, involving a combination of various autoantibodies and other factors rather than a single causative factor. First, genome-wide association studies have not identified a single gene mutation strongly linked to SSc.^42^ Second, the disease concordance in twins is modest and similar between monozygotic and dizygotic twins, further supporting the notion that SSc is not caused only by genetic factors.^43^ Finally, and most importantly, the clinical manifestations of SSc are highly heterogeneous.^1^

Alternatively, a cluster of several autoantibodies, with combinations that vary among patients, might explain the heterogeneity of clinical manifestations in SSc.^44^ Co-localization of ATA and candidate autoantibodies within the same module identified by WGCNA, as well as the distribution of ATA, ARA, and ACA among different modules (**Figure 4**), support this hypothesis. Such clustering of autoantibodies might result from cross reaction with a single specific exogenous antigen such as viruses,^45,46^ or intermolecular epitope spreading, a phenomenon we have recently demonstrated to be involved in SSc progression using the same WPA system.^47^ Further research is needed to unravel the relationships among these autoantibodies and their collective contribution to SSc development. This research should include comparisons of structural similarity and epitope mapping for the relevant antigens, and longitudinal serological monitoring in each case. Understanding these interactions will be crucial for comprehensively elucidating the mechanisms underlying the autoimmune aspect of SSc and potentially identifying new therapeutic targets.

Our study has several notable strengths. Firstly, the data presented is derived from a prospective, randomized, placebo-controlled clinical trial, which includes two time points (before and after RTX therapy). This adherence to good clinical practice ensures high data accuracy. Secondly, the use of the wheat-germ *in vitro* protein synthesis system and the manipulation technique for WPAs enabled high-throughput expression of a variety of human proteins, including membranous proteins, on a single platform.^18^ As a result, our autoantibody measurement could encompass a wider range of antigens at an almost proteome-wide level, allowing for the application of omics-based bioinformatics approaches to interpret the data.^24^ Meanwhile, one major limitation is the lack of functional assays both *in vitro* and *in vivo*. Further investigation is needed to determine whether the candidate autoantibodies we identified act as agonists or antagonists to their target proteins, and whether they influence the pathophysiology of SSc. Moreover, the value of autoantibodies we identified for predicting treatment response to RTX should be validated in external cohorts in real-world clinical settings.

## Data Availability

All data produced in the present study are available upon reasonable request to the authors.

## Acknowledgements

We thank Ms. Maiko Enomoto and her colleagues for their secretarial work. We appreciate K. Yamaguchi, T. Okumura, C. Ono, A. Sato, A. Miya, and N. Goshima from ProteoBridge Corporation for preparing the WPAs. We also acknowledge R. Uchino, Y. Murakami, and H. Matsunaka from TOKIWA Pharmaceuticals Co. Ltd. for providing technical assistance with autoantibody measurement.

## Author Contributions

KM Matsuda primarily engaged in autoantibody measurement, clinical data collection, data analysis, visualization, and writing the first draft of the manuscript. K Iwadoh participated in machine learning analysis. S Ebata was primarily engaged in the management of SSc patients participated in the DESIRES trial. H Kotani, A Kuzumi, T Fukasawa, A Yoshizaki-Ogawa took part in the sample collection of SSc. S Sato conceptualized and supervised the study. A Yoshizaki conceptualized, launched, and supervised this study, and was involved in revising the manuscript.

## Methods

### Study Design

The study design for the DESIRES trial has been previously reported (NCT04274257).^13^ The full protocol of the trial is available at https://clinicaltrials.gov/study/NCT04274257. Briefly, the DESIRES trial was a randomized, double-blind, placebo-controlled trial of 24 weeks. The primary endpoint of the double-blind phase was the absolute change in mRSS 24 weeks after 13 intervention initiation compared to the baseline. In total, 56 patients were randomized to receive either intravenous rituximab (375 mg/m^2^) or matching placebo once per week for 4 weeks, based on the allocation factors of 4 disease duration (≤6 years or >6 years), mRSS (≥20 or <20), and concomitant ILD (present or absent) by the minimization method. Of these, 49 patients completed the double-blind phase. Serum samples were collected and served for PWAS at the beginning and the end of the double-blind phase (**Fig. 1A**). After excluding 4 cases due to their serum unsuitable for PWAS, data of 45 patients were included in our analysis. Age and sex matched HCs were selected from a cohort of healthcare providers on annual checkups without any medical history (**Fig. 1B**), using “matchIt” R package.^48^ This study was approved by the ethics committee of the University of Tokyo Graduate School of Medicine and conducted in accordance with the Declaration of Helsinki. Written informed consent was obtained from all patients.

### Assessments

Clinical and laboratory assessments were performed at baseline and at 24 weeks after the first infusion of rituximab. Skin sclerosis was assessed by mRSS.^49^ Lung function was evaluated by pulmonary function tests. Serum levels of Krebs von den Lungen-6 (KL-6; normal range: 0-500 U/mL) and surfactant protein-D (SP-D; normal range: 0-110 ng/mL), which are glycoproteins mainly produced by type II pneumocytes, were measured as established markers of ILD in patients with SSc. Laboratory examinations included white blood cell count, lymphocyte count, the number of CD19^+^ and CD20^+^ cells, and serum levels of IgG (normal range: 700-1600 mg/dL), IgM (normal range: 40-250 mg/dL), and IgA (normal range: 70-400 mg/dL). Patients with mRSS improvement of 7 or higher and 6 or lower were classified as HRs and LRs, respectively, based on the previous study on minimally important differences for mRSS.^50^ This classification resulted in 16 high responders and 13 LRs (**Fig. 1C**).

### Autoantibody measurement

WPAs were arranged as previously described.^20^ First, proteins were synthesized *in vitro* utilizing a wheat germ cell-free system from 13,455 clones of the HuPEX.^18^ Second, synthesized proteins were plotted onto glass plates (Matsunami Glass, Osaka, Japan) in an array format by the affinity between the GST-tag added to the N-terminus of each protein and glutathione modified on the plates. The WPAs were treated with human serum diluted by 3:1000 in the reaction buffer containing 1x Synthetic block (Invitrogen), phosphate-buffered saline (PBS), and 0.1% Tween 20. Next, the WPAs were washed, and goat anti-Human IgG (H+L) Alexa Flour 647 conjugate (Thermo Fisher Scientific, San Jose, CA, USA) diluted 1000-fold was added to the WPAs and reacted for 1 hour at room temperature. Finally, the WPAs were washed, air-dried, and fluorescent images were acquired using a fluorescence imager (Typhoon FLA 9500, Cytiva, Marlborough, MA, USA). Fluorescence images were analyzed to quantify serum levels of autoantibodies targeting each antigen, following the formula shown below:

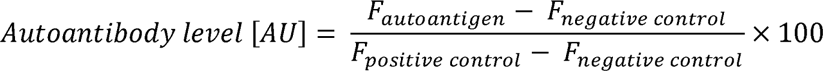

*AU*: arbitrary unit
*F _autoantigen_*: fluorescent intensity of autoantigen spot
*F _negative control_*: fluorescent intensity of negative control spot
*F _positive control_*: fluorescent intensity of positive control spot

### Machine learning

We applied supervised machine learning techniques using the Python code with the scikit-learn library to analyze the autoantibody measurement data. With the random forest model, decision trees were built and trained in parallel on subsets of sampled instances and features. Meanwhile, with the XGBoost model, decision trees were built sequentially to improve each other. The final prediction of the random forest was based on the majority of its decision trees, while that of XGBoost was derived from their weighted average. The performance of the classifiers was evaluated using the area under the operator-receiver characteristics curve (AUC), accuracy, precision, recall, and F1-score, with the higher the scores indicating the better classification performance. The accuracy is the ratio of the correct positive and negative prediction, the precision is the ratio of the correct positive prediction, the recall (or sensitivity) is the ratio of the correct positive prediction among all true positive instances, and F1-score is the harmonic mean of precision and sensitivity.

### WGCNA analysis

The weighted gene co-expression network was constructed using the “WGCNA” R package.^28^ We calculated each gene pair’s Pearson correlation coefficient, measured how similarly their expressions were expressed, and created a correlation matrix. Scale-free topology requirements were used to compute the “soft” threshold power to build biologically meaningful scale-free networks. Based on the adjacency matrix, dynamic tree cuts and at least 100 genes per module were utilized to generate a topological overlap matrix for co-expression modules. In addition, we assessed gene significance, module membership, and correlated modules with clinical characteristics and mapped signature genes.

### Statistical analysis

Fisher exact test was performed to compare categorical variables. Mann-Whitney U test or Wilcoxon signed-rank test as appropriate was performed to compare continuous variables. Spearman correlation test was used for correlation analysis. P values of < 0.05 were considered statistically significant. Gene Ontology Analysis using web-based tools targeted the list of the entry clones coding the differentially highlighted autoantigens was performed for gene-list enrichment analysis, gene-disease association analysis, and transcriptional regulatory network analysis with Metascape.^25^ Data analyses were conducted using R (v4.2.1).

### Data visualization

Box plots, scatter plots, hierarchical clustering, and correlation matrix were visualized by using R (v4.2.1). Box plots were defined as follows: the middle line corresponds to the median; the lower and upper hinges correspond to the first and third quartiles; the upper whisker extends from the hinge to the largest value no further than 1.5 times the interquartile range (IQR) from the hinge; and the lower whisker extends from the hinge to the smallest value at most 1.5 times the IQR of the hinge.

**Extended Figure 1. Additional results of PWAS. (A)** The sum of autoantibody levels (SAL) by age groups. **(B)** Serum levels of SSc-related autoantibodies before and after the DESIRES trial by the treatment arm and responsiveness.

**Extended Figure 2. Autoantibodies highlighted in each machine learning model. (A)** Autoantibodies that were mostly highlighted according to feature importance by Lasso regression, Ridge regression, SVM with normalization, Random Forest, XGBoost, and LightGBM. **(B)** The inclusion relationship of autoantibodies highlighted by the six machine learning frameworks illustrated by an UpSet plot. **(C)** The box plots describe the serum levels of autoantibodies highlighted by more than two frameworks in COVID-19, atopic dermatitis (AD), anti-neutrophil cytoplasmic antibody-associated vasculitis (AAV), systemic lupus erythematosus (SLE), systemic sclerosis (SSc), and healthy controls (HCs). The data derives from the UT-ABCD.

**Extended Figure 3. Distribution of the candidate autoantibodies among various disorders.** The box plots describe the serum levels of the candidate autoantibodies associated with “import across plasma membrane” **(A)** and “peptide GPCRs” **(B)** in COVID-19, atopic dermatitis (AD), anti-neutrophil cytoplasmic antibody-associated vasculitis (AAV), systemic lupus erythematosus (SLE), systemic sclerosis (SSc), and healthy controls (HCs). The data derives from the UT-ABCD.

**Extended Figure 4. Expression of highlighted autoantigens in human tissues and single cells.** Expression of autoantigens associated with “import across plasma membrane” **(A)** or with “peptide GPCRs” **(B)** in multiple human tissues, as measured by bulk RNA-sequencing from the Human Protein Atlas. Expression of CCR8 **(C)** or FPR1 **(D)** in immune cells from multiple human tissues, as measured by single-cell RNA-sequencing from the Tabula Sapiens project.

