## Supplementary figures and images for "Deciphering autoantibody landscape of systemic sclerosis through systems-based approach: insights from a B-cell depletion clinical trial"

### RTX extfig1.tiff

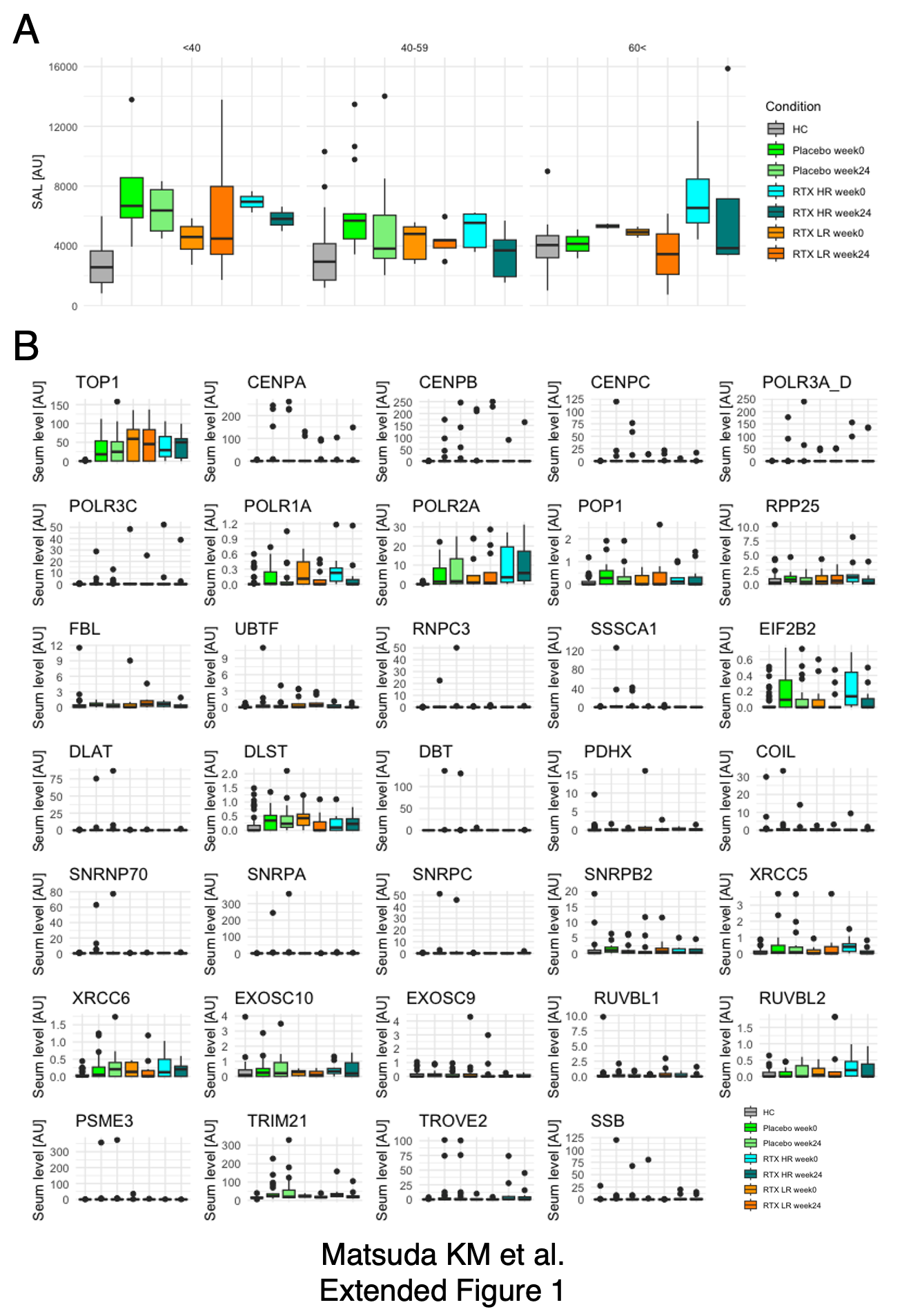

### RTX extfig2.tiff

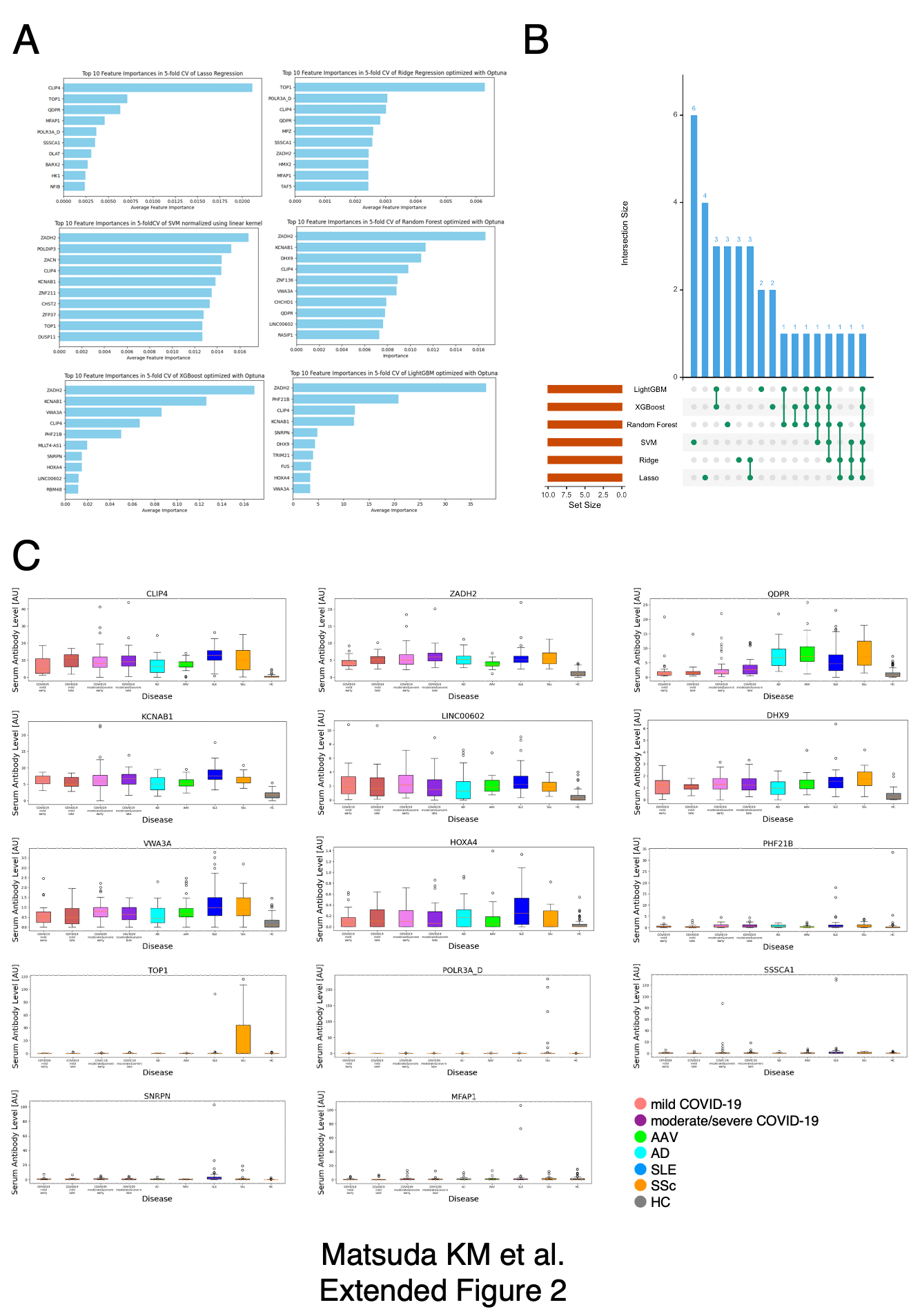

### RTX extfig3.tiff

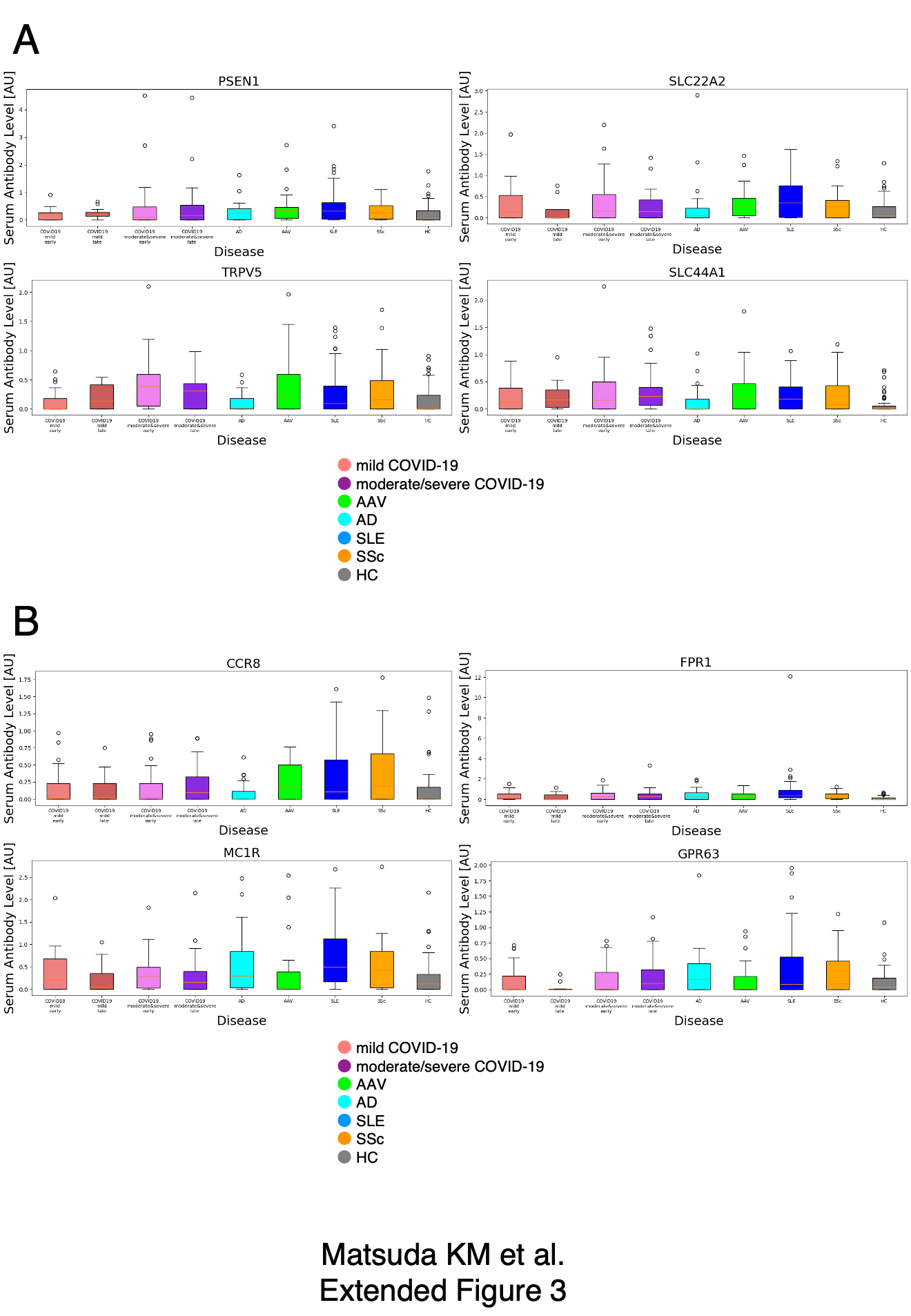

### RTX extfig4.tiff

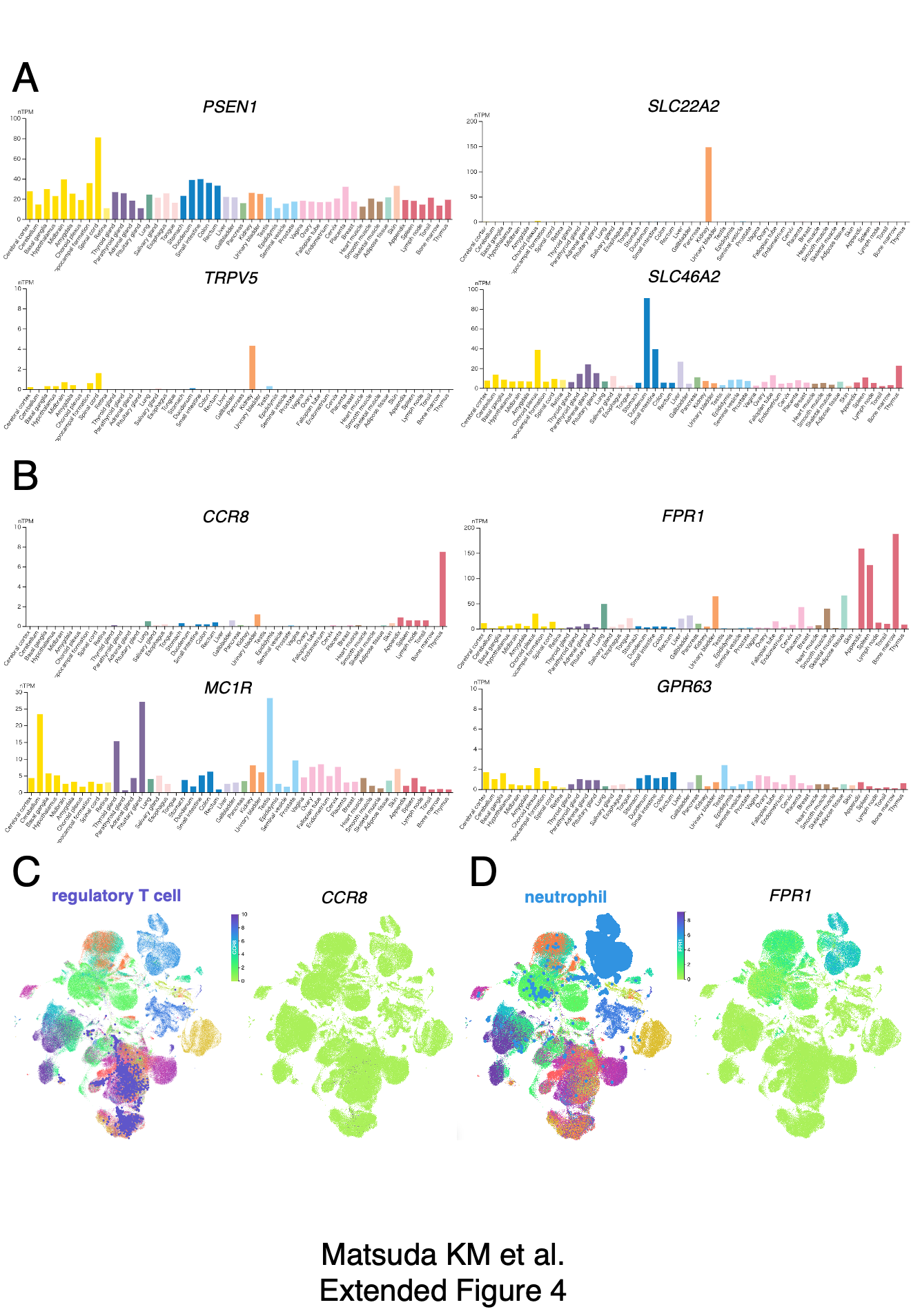
